# Comparison of Multimorbidity in COVID-19 infected and general population in Portugal

**DOI:** 10.1101/2020.07.02.20144378

**Authors:** Miguel Froes, Bernardo Neves, Bruno Martins, Mário J. Silva

## Abstract

Understanding COVID-19 and its risk factors in the Portuguese population is critical to the struggle against this infectious disease. To study the impact of multimorbidity in the population with COVID-19 infection, we performed a descriptive analysis of a dataset extracted from all reported confirmed cases of COVID-19 in Portugal until June 30, 2020. We observed a prevalence of multimorbidity in 6.77% of the 36,244 infected patients. These patients showed an increased risk of hospitalization, ICU admission, and mortality with OR 2.22 (CI 95%: 2.13-2.32) for every additional morbidity. Further studies should confirm these findings and special attention should be made on data collection, to ensure proper recording of patient comorbidities.

## 1 Introduction

COVID-19 is an infectious disease caused by SARS-CoV-2 that recently started challenging health systems. Its impact is global with 21,200,000 cases and 761,000 deaths by August 16, 2020 [1]. Early evidence from the pandemic suggested that older patients with chronic conditions were over-represented and may herald a poor clinical course. However, data supporting these analysis is still largely limited to China and Italy [2, 3]. Given that multimorbidity increases with age, the specific risk of different chronic conditions and their combination in terms of poor health outcomes needs to be adjusted for age, which is not routinely done [4]. Understanding which groups are at higher risk is important to better inform public health policies and resource allocation, and to advance knowledge about this novel condition.

DGS (Directionate-General of Health), the Portuguese public health authority, developed and operates SINAVE (National Epidemiological Surveillance System), i.e. the national public health surveillance system. SINAVE is used to collect, update, analyze, and disclose data related to infectious diseases with mandatory reporting, and other public health hazards [5]. Symptoms, previous medical history, disease course, and laboratory results are introduced by physicians in charge of COVID-19 patients. Although SINAVE was not specifically designed for this outbreak, it has been used since the beginning of the pandemic as the main official source of data about COVID-19 cases in Portugal.

In this work, we evaluate the prevalence of multimorbidity and age-adjusted risk of hospitalization, ICU admission, and death, in the Portuguese population from official data, based on a dataset^2^ extracted from SINAVE containing all confirmed cases of COVID-19 infection, in Portugal, by June 30, 2020. Our analysis is based on the DGS/SINAVE dataset spanning the full period (i.e., the June version of the dataset, which is the most recent at the time of this report), which updates an initial (April) version of the dataset based on cases reported until April 28, 2020, adding two more months and providing more data about the initial cases.

## 2 Methods

### 2.1 Sample

Our retrospective observational study used data provided by DGS after the required institutional and ethical approvals. The sample population consists of all the Portuguese population with SARS-CoV2 confirmed infection as notified by clinicians by June 30, 2020. A broad range of clinical and demographic variables are present in this dataset. In our study, we specifically used variables corresponding to age, gender, hospital admission, admission in intensive care unit, mortality, and patient’s underlying conditions.

### 2.2 Measures

Chronic conditions were originally provided in the dataset as categorical variables on the presence, absence, or unknown status of the following conditions: asthma, malignancy, chronic hematological disorder, diabetes, HIV/other immune deficiency, renal disease, liver disease, chronic lung disease, and neuromuscular disorder.

A field containing *raw* textual input from doctors was also taken into account, to better complement the cases where the chronic conditions were left as unknown. This alternative information was indeed very useful, particularly on what regards cardiac disorders (including hypertension and other cardiovascular diseases), which were not included in the dataset as a categorical variable and could, therefore, not be detected if not for the *raw* input.

A text mining script, using keywords associated with all the previously mentioned conditions, was used in order to better capture the prevalence of the diseases (i.e., in many cases, we noticed that although the categorical variables corresponding to chronic conditions were left with an unknown status, the textual inputs contained relevant mentions to chronic conditions) and to detect cases of cardiac disorders. The following keywords (in Portuguese) were used, in connection to each of the diseases that was considered in our study:

- **Asthma:** asma
- **Malignancy:** neo, cancro, carcinoma, linfoma
- **Cardiac disorder (including hypertension and other cardiovascular diseases):** cardio, cárdio, miocar, cardía, cardia, hta, auricular, arterial, venosa
- **Chronic hematological disorder:** hematológica
- **Diabetes:** diabetes, DM
- **HIV/other immune deficiency:** hiv, vih
- **Renal disease:** renal
- **Liver disease:** hepatomegalia
- **Chronic lung disease:** dpoc, pulmonar
- **Neuromuscular disorder:** alz, parkinson, epilepsia.

The keywords were chosen in order to cover different cases, considering misspellings or abbreviations. For instance, *neoplasia* was commonly abbreviated to *neo* and the latter was therefore preferred (and also matched as a prefix). Another example is the use of the prefix *alz* to detect Alzheimer’s disease, that was often misspelled as *Alzaimer*.

The contribution of using the *raw* textual input when, preparing the data, was also analyzed. All the statistics reported on this article were also measured in a version of the data that only considered the structured information. The main difference, as stated above, was the complete absence of any information on cardiac disorders, when not considering the textual information. There were also some differences in the prevalence of the other chronic conditions, although only reaching up to 0.16%.

We defined multimorbidity as the presence of two or more conditions in the same individual, following the definition used by other authors [6]. As will be further discussed on the next section, multimorbidity was present on 6.77% of the cases in the dataset, when considering the textual inputs, while this number would be reduced to 4.01% if only considering the structured information. Addressed outcomes were hospitalization, admission to ICU unit, and reported death. A composite outcome of any of these events was also analyzed.

### 2.3 Statistical analysis

We used *Python 3.6* and packages *Numpy, Pandas, Seaborn*, and *SciPy*, in combination with *Microsoft Excel* to evaluate and plot the prevalence of multimorbidity. Additionally, *IBM SPSS Statistics* was used for obtaining age-adjusted risk of hospitalization, ICU admission, and death.

Categorical variables were presented to analysis as counts and percentages with 95% confidence intervals, and comparisons were made using using the χ^2^ test. Univariate regression analysis of each individual chronic condition was performed adjusting only for age. A multivariate logistic regression was performed adjusting for age and every other chronic condition with significant statistical association on univariate analysis. Results were considered statistically significant when P <0,05.

## 3 Results

The overall sample contained 36,244 adult patient cases, with women being more prevalent (56.66%). Among the cases, 18.79% had at least one chronic condition. Cardiac disorder was the most commonly reported condition, respectively in 43.33% of the patients with any morbidity. Table 1 shows the reported prevalence of different chronic conditions in the studied population. Multimorbidity, as previously defined, was present in 6.77% of the cases. Figures 1 and 2 plot the prevalence of multimorbidity by age group, respectively for (a) the COVID-19 infected general population, and for (b) the hospitalized population. To analyze the Odd Ratio and prevalence of co-occurring pairs of chronic diseases, people with unknown disease prevalence were excluded, which resulted in a population of 33,283 adult patients. The prevalence of co-occurring pairs of chronic health conditions, plotted in Figure 4, shows Cardiac disorders and Diabetes as the most common dyad of chronic diseases.

**Table 1:**
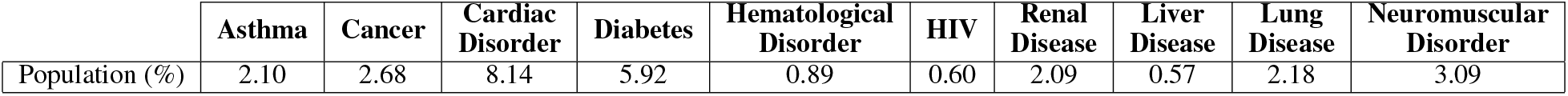
Percentage of COVID-19 infected total Portuguese population affected by each comorbidity.

|  | **Asthma** | **Cancer** | **Cardiac Disorder** | **Diabetes** | **Hematological Disorder** | **HIV** | **Renal Disease** | **Liver Disease** | **Lung Disease** | **Neuromuscular Disorder** |
| --- | --- | --- | --- | --- | --- | --- | --- | --- | --- | --- |
| Population (%) | 2.10 | 2.68 | 8.14 | 5.92 | 0.89 | 0.60 | 2.09 | 0.57 | 2.18 | 3.09 |

**Table 2:** Odds Ratio for the outcome (Death) for the Age variable and analyzed comorbidities. The real value of Age was used instead of an age group. Being a continuous variable, an Odds Ratio of 1.097 implies that, as the Age variable increases by 1, the probability of the patient having the outcome increases by 9.7%. **B**, **S.E**, and **Wald** are the unstandardized regression weight, how much the unstandardized regression weight can vary by, and test statistic for the individual predictor variable, respectively.

|  | **B** | **S.E.** | **Wald** | **df** | **Sig.** | **Exp(B)** | **95% C.I.** |  |
| --- | --- | --- | --- | --- | --- | --- | --- | --- |
|  |  |  |  |  |  |  | **Lower** | **Upper** |
| **Age** | ,092 | ,002 | 1446,655 | 1 | <0,001 | 1,097 | 1,092 | 1,102 |
| **Asthma** | ,188 | ,262 | ,515 | 1 | ,473 | 1,207 | ,722 | 2,017 |
| **Cancer** | 1,021 | ,113 | 82,084 | 1 | <0,001 | 2,775 | 2,225 | 3,461 |
| **Cardiac Disorder** | ,520 | ,081 | 41,384 | 1 | <0,001 | 1,682 | 1,435 | 1,970 |
| **Diabetes** | ,723 | ,087 | 69,373 | 1 | <0,001 | 2,061 | 1,739 | 2,444 |
| **Hematologic Disease** | ,997 | ,175 | 32,438 | 1 | <0,001 | 2,711 | 1,924 | 3,822 |
| **HIV** | 1,330 | ,306 | 18,837 | 1 | <0,001 | 3,780 | 2,074 | 6,892 |
| **Renal Disease** | 1,177 | ,108 | 118,925 | 1 | <0,001 | 3,244 | 2,626 | 4,008 |
| **Liver Disease** | 1,437 | ,253 | 32,137 | 1 | <0,001 | 4,207 | 2,560 | 6,914 |
| **Lung Disease** | ,897 | ,117 | 58,276 | 1 | <0,001 | 2,452 | 1,948 | 3,087 |
| **Neuromuscular Disease** | 1,092 | ,091 | 143,005 | 1 | <0,001 | 2,981 | 2,492 | 3,565 |

**Table 3:** Odds Ratio for the outcome (Hospitalization) for the Age variable and analyzed comorbidities. The real value of Age was used instead of an age group. Being a continuous variable, an Odds Ratio of 1.056 implies that, as the Age variable increases by 1, the probability of the patient having the outcome increases by 5.6%. **B**, **S.E**, and **Wald** are the unstandardized regression weight, how much the unstandardized regression weight can vary by, and test statistic for the individual predictor variable, respectively.

|  | **B** | **S.E.** | **Wald** | **df** | **Sig.** | **Exp(B)** | **95% C.I** |  |
| --- | --- | --- | --- | --- | --- | --- | --- | --- |
|  |  |  |  |  |  |  | **Lower** | **Upper** |
| **Age** | ,054 | ,001 | 3180,583 | 1 | <0,001 | 1,056 | 1,054 | 1,058 |
| **Asthma** | ,085 | ,130 | ,429 | 1 | ,513 | 1,088 | ,844 | 1,403 |
| **Cancer** | 1,251 | ,076 | 271,427 | 1 | <0,001 | 3,493 | 3,010 | 4,054 |
| **Cardiac Disorder** | ,815 | ,049 | 277,596 | 1 | <0,001 | 2,258 | 2,052 | 2,486 |
| **Diabetes** | 1,205 | ,054 | 494,291 | 1 | <0,001 | 3,335 | 2,999 | 3,709 |
| **Hematologic Disease** | 1,772 | ,144 | 151,367 | 1 | <0,001 | 5,882 | 4,435 | 7,801 |
| **HIV** | 1,538 | ,158 | 94,268 | 1 | <0,001 | 4,654 | 3,412 | 6,348 |
| **Renal Disease** | 2<0,001 | ,091 | 478,288 | 1 | <0,001 | 7,387 | 6,175 | 8,836 |
| **Liver Disease** | 1,885 | ,166 | 128,229 | 1 | <0,001 | 6,584 | 4,751 | 9,123 |
| **Lung Disease** | 1,307 | ,086 | 232,600 | 1 | <0,001 | 3,694 | 3,123 | 4,369 |
| **Neuromuscular Disease** | 1,536 | ,076 | 409,383 | 1 | <0,001 | 4,645 | 4,003 | 5,390 |

**Table 4:**
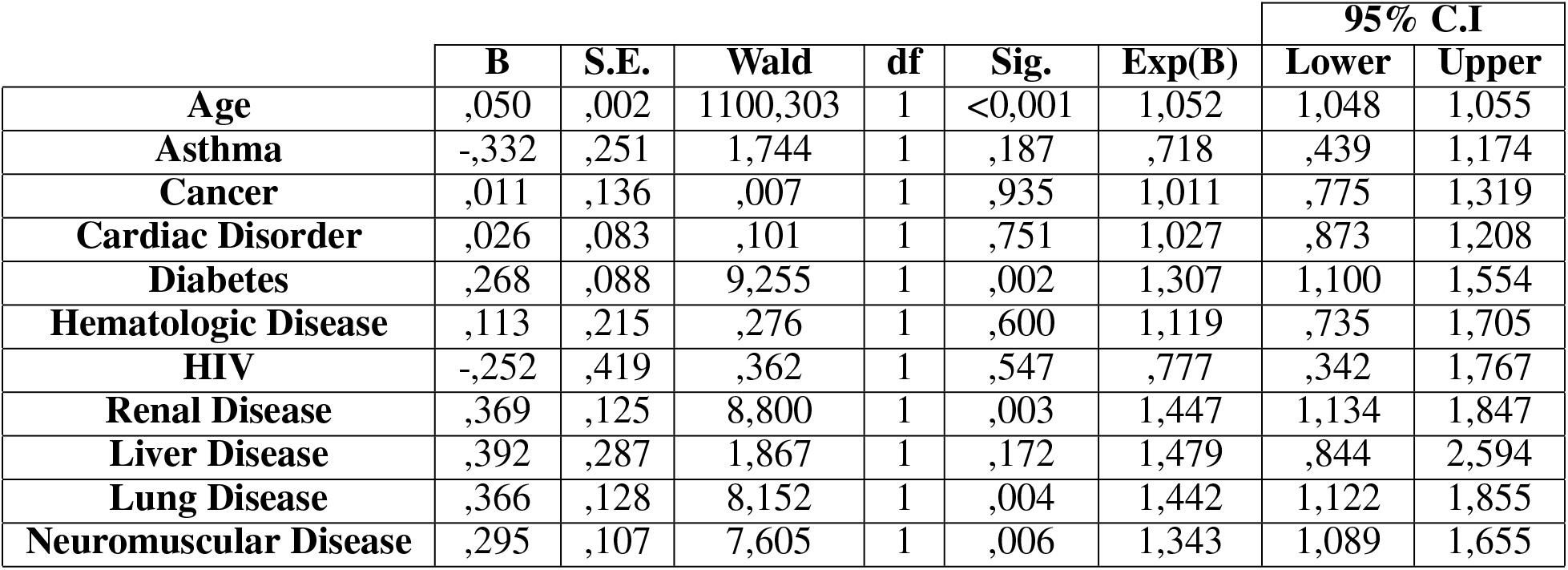
Odds Ratio for the outcome (ICU stay) for the Age variable and analyzed comorbidities. The real value of Age was used instead of an age group. Being a continuous variable, an Odds Ratio of 1.052 implies that, as the Age variable increases by 1, the probability of the patient having the outcome increases by 5.2. **B**, **S.E**, and **Wald** are the unstandardized regression weight, how much the unstandardized regression weight can vary by, and test statistic for the individual predictor variable, respectively.

|  | **B** | **S.E.** | **Wald** | **df** | **Sig.** | **Exp(B)** | **95% C.I** |  |
| --- | --- | --- | --- | --- | --- | --- | --- | --- |
|  |  |  |  |  |  |  | **Lower** | **Upper** |
| **Age** | ,050 | ,002 | 1100,303 | 1 | <0,001 | 1,052 | 1,048 | 1,055 |
| **Asthma** | -,332 | ,251 | 1,744 | 1 | ,187 | ,718 | ,439 | 1,174 |
| **Cancer** | ,011 | ,136 | ,007 | 1 | ,935 | 1,011 | ,775 | 1,319 |
| **Cardiac Disorder** | ,026 | ,083 | ,101 | 1 | ,751 | 1,027 | ,873 | 1,208 |
| **Diabetes** | ,268 | ,088 | 9,255 | 1 | ,002 | 1,307 | 1,100 | 1,554 |
| **Hematologic Disease** | ,113 | ,215 | ,276 | 1 | ,600 | 1,119 | ,735 | 1,705 |
| **HIV** | -,252 | ,419 | ,362 | 1 | ,547 | ,777 | ,342 | 1,767 |
| **Renal Disease** | ,369 | ,125 | 8,800 | 1 | ,003 | 1,447 | 1,134 | 1,847 |
| **Liver Disease** | ,392 | ,287 | 1,867 | 1 | ,172 | 1,479 | ,844 | 2,594 |
| **Lung Disease** | ,366 | ,128 | 8,152 | 1 | ,004 | 1,442 | 1,122 | 1,855 |
| **Neuromuscular Disease** | ,295 | ,107 | 7,605 | 1 | ,006 | 1,343 | 1,089 | 1,655 |

**Table 5:** Odds Ratio for the outcome (Death + hospitalization + ICU stay) for the Age variable and analyzed comorbidities. The real value of Age was used instead of an age group. Being a continuous variable, an Odds Ratio of 1.061 implies that, as the Age variable increases by 1, the probability of the patient having the outcome increases by 6.1%. **B**, **S.E**, and **Wald** are the unstandardized regression weight, how much the unstandardized regression weight can vary by, and test statistic for the individual predictor variable, respectively.

|  | **B** | **S.E.** | **Wald** | **df** | **Sig.** | **Exp(B)** | **95% C.I** |  |
| --- | --- | --- | --- | --- | --- | --- | --- | --- |
|  |  |  |  |  |  |  | **Lower** | **Upper** |
| **Age** | ,059 | ,001 | 3654,861 | 1 | <0,001 | 1,061 | 1,059 | 1,063 |
| **Asthma** | ,032 | ,130 | ,060 | 1 | ,806 | 1,033 | ,800 | 1,333 |
| **Cancer** | 1,216 | ,076 | 253,311 | 1 | <0,001 | 3,373 | 2,904 | 3,918 |
| **Cardiac Disorder** | ,768 | ,049 | 248,499 | 1 | <0,001 | 2,156 | 1,959 | 2,372 |
| **Diabetes** | 1,170 | ,054 | 464,558 | 1 | <0,001 | 3,221 | 2,896 | 3,583 |
| **Hematologic Disease** | 1,747 | ,147 | 140,286 | 1 | <0,001 | 5,736 | 4,296 | 7,658 |
| **HIV** | 1,537 | ,159 | 93,498 | 1 | <0,001 | 4,649 | 3,405 | 6,348 |
| **Renal Disease** | 2,032 | ,094 | 463,142 | 1 | <0,001 | 7,627 | 6,338 | 9,177 |
| **Liver Disease** | 1,811 | ,168 | 116,114 | 1 | <0,001 | 6,115 | 4,399 | 8,501 |
| **Lung Disease** | 1,320 | ,087 | 231,704 | 1 | <0,001 | 3,743 | 3,158 | 4,437 |
| **Neuromuscular Disease** | 1,586 | ,078 | 414,126 | 1 | <0,001 | 4,886 | 4,194 | 5,693 |

**Figure 1:**
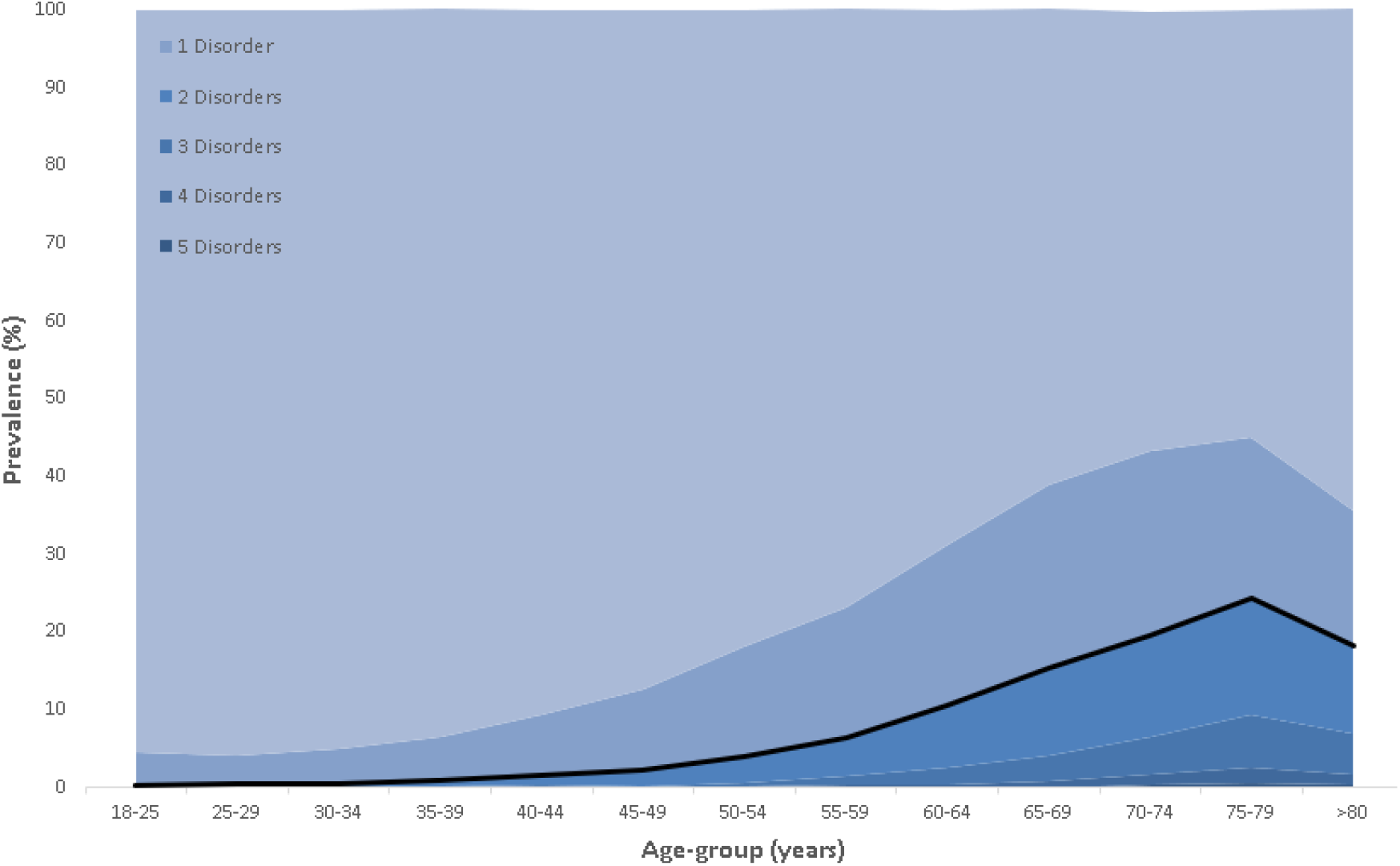
Prevalence of multimorbidity by age group for the COVID-19 infected Portuguese population. The lighter shade of blue is representative of the absence of conditions and the black line represents the prevalence of multimorbidity.

**Figure 2:**
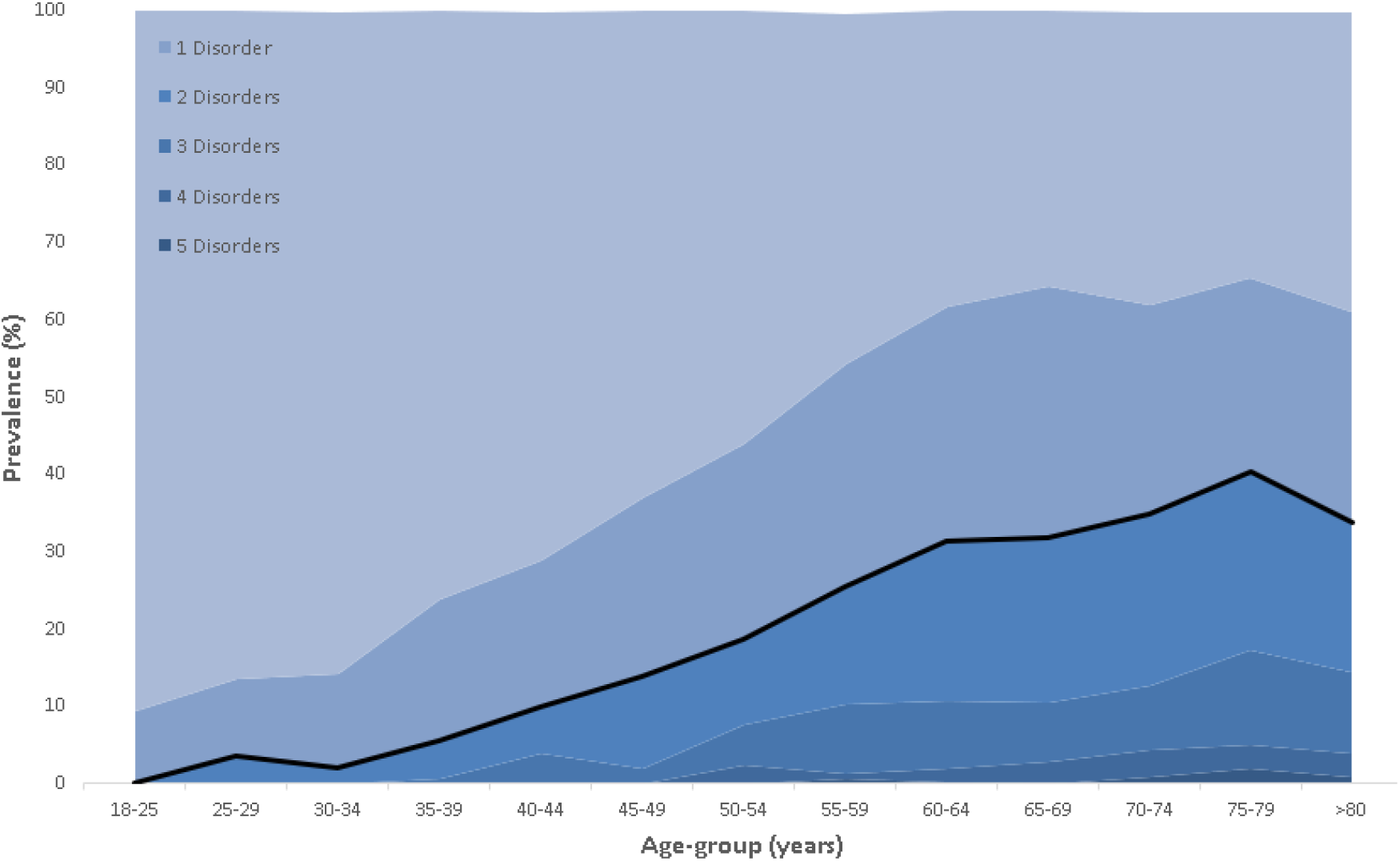
Prevalence of multimorbidity by age group for the COVID-19 infected Portuguese hospitalized population. The lighter shade of blue is representative of the absence of conditions and the black line represents the prevalence of multimorbidity.

**Figure 3:**
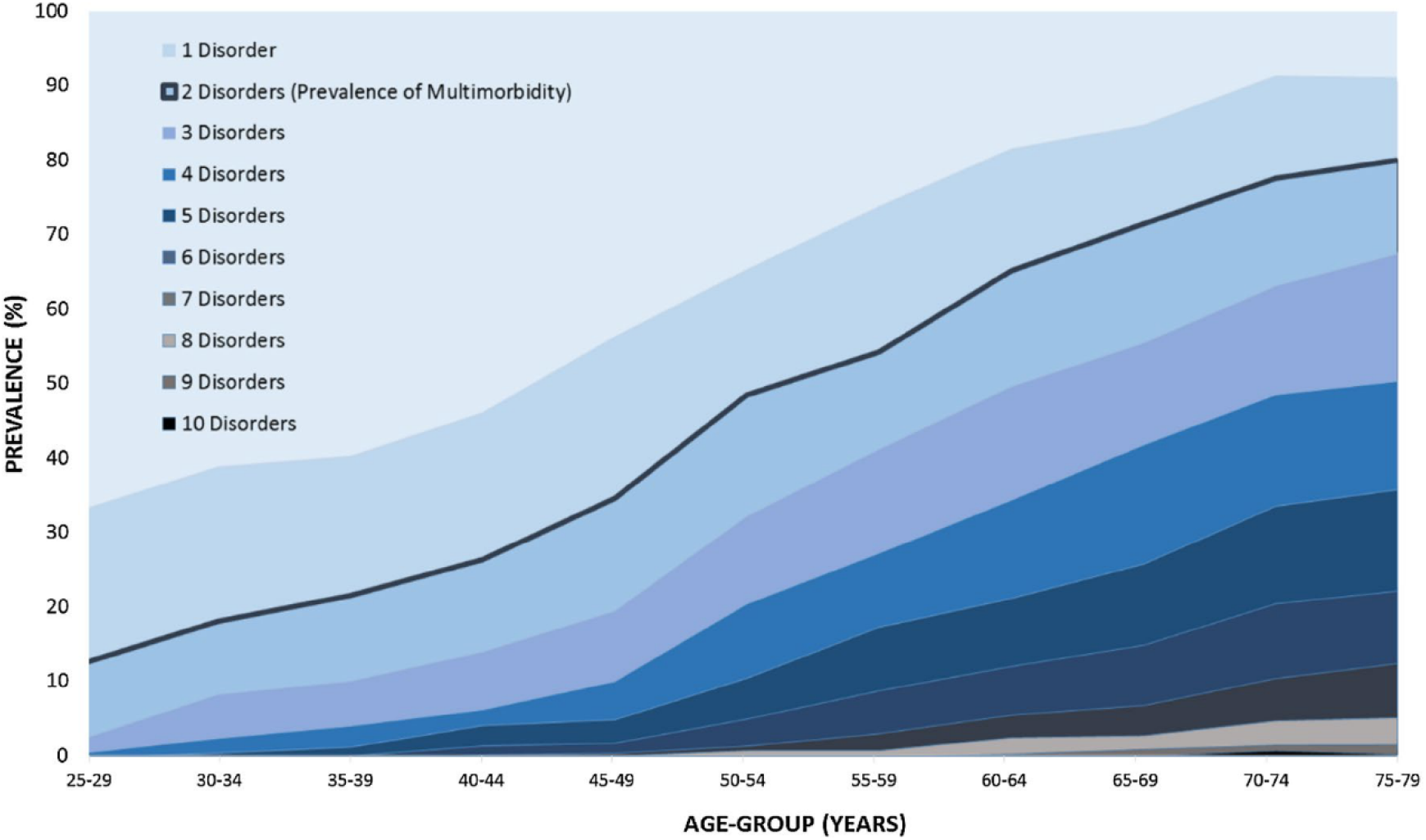
Prevalence of multimorbidity by age group using data from the Portuguese Fifth National Health Interview Survey (Laires et al. 2019). The lighter shade of blue is representative of the absence of conditions and the black line represents the prevalence of multimorbidity.

**Figure 4:**
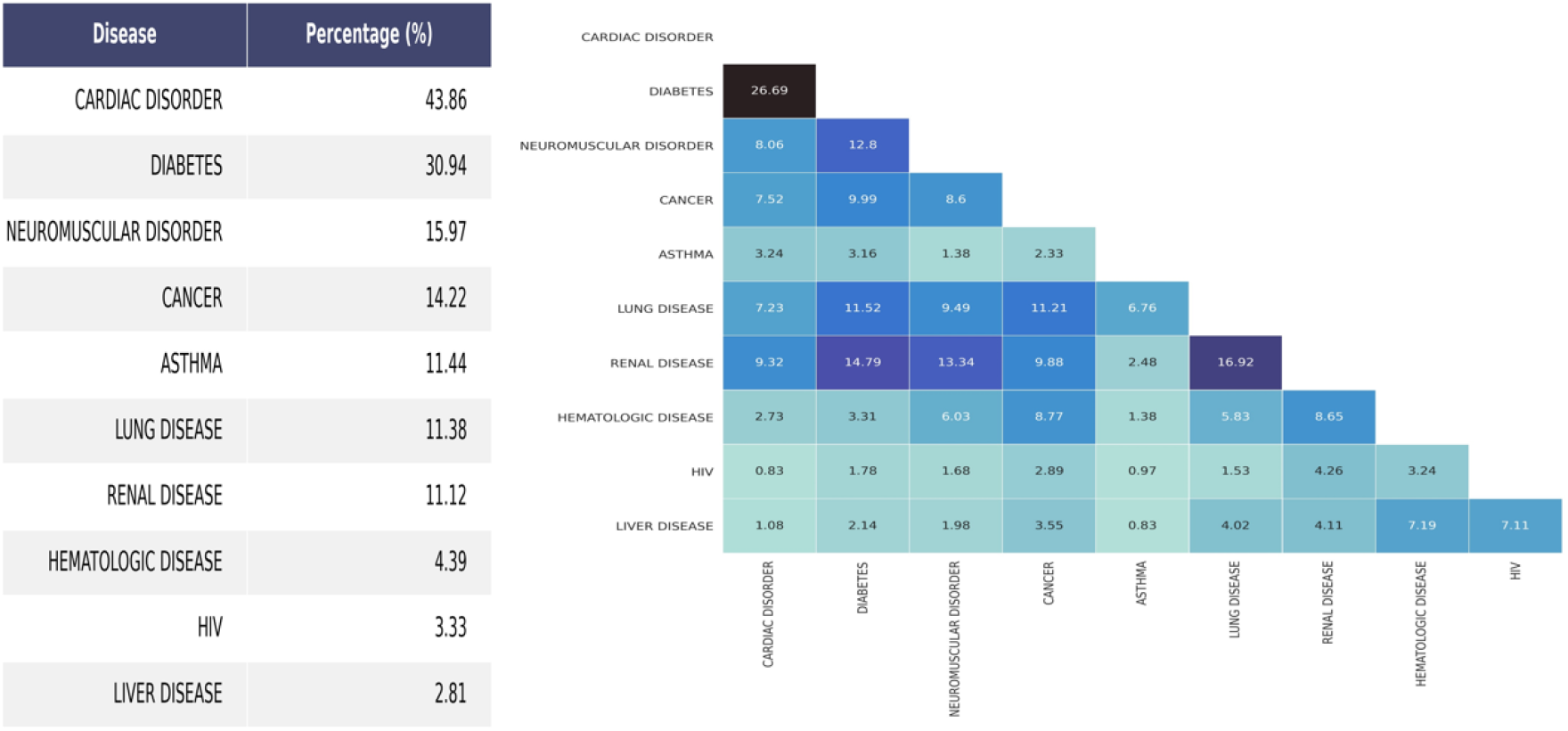
Prevalence of single (left) and co-occurring pairs (right) of chronic health conditions. Left: Prevalence of the disease in the Portuguese population with at least one disease. Right: Prevalence of the disease, rows, in the Portuguese population affected by another disease, columns. All values are presented in percentage.

Data regarding hospitalization and ICU admission was available for only 32,945 patients (90.90% of the overall study population). Within this population, hospitalization occurred in 12.89% of the patients, with a male predominance (50.66%), and ICU admission was required for 4.11% of the patients, with a female predominance (51.73%). Observed mortality was 3.19%. All chronic conditions, except for asthma, were associated with increased risk of mortality and hospitalization (Tables 6 and 7, respectively). Age, diabetes, renal disease, lung disease, and neuromuscular disorders, were all associated with increased risk of ICU admission (Table 8). Additionally, every additional chronic condition increases the risk for the patients of the composite outcome of death, hospitalization, or ICU admission, by 123.3% (OR 2.22; CI95%: 2.13-2.32).

**Table 6:** Odds Ratio for the association between the age, categories of comorbidity and outcome (Death) in patients with COVID-19. The continuous value of Age was used instead of an age group. Being a continuous variable, an Odds Ratio of 1.093 implies that, as the Age variable increases by 1, the probability of the patient having the outcome increases by 9.3%. **B**, **S.E**, and **Wald** are the unstandardized regression weight, how much the unstandardized regression weight can vary by, and test statistic for the individual predictor variable, respectively.

|  | **B** | **S.E.** | **Wald** | **df** | **Sig.** | **Exp(B)** | **95% C.I** |  |
| --- | --- | --- | --- | --- | --- | --- | --- | --- |
|  |  |  |  |  |  |  | **Lower** | **Upper** |
| **Age** | ,089 | ,003 | 1157,213 | 1 | <0,001 | 1,093 | 1,088 | 1,099 |
| **Cancer** | ,711 | ,119 | 35,639 | 1 | <0,001 | 2,037 | 1,613 | 2,573 |
| **Cardiac Disorder** | ,257 | ,086 | 8,903 | 1 | ,003 | 1,293 | 1,092 | 1,530 |
| **Diabetes** | ,253 | ,096 | 6,887 | 1 | ,009 | 1,288 | 1,066 | 1,556 |
| **Hematologic Disease** | ,253 | ,186 | 1,864 | 1 | ,172 | 1,288 | ,896 | 1,854 |
| **HIV** | ,890 | ,320 | 7,750 | 1 | ,005 | 2,436 | 1,301 | 4,558 |
| **Renal Disease** | ,698 | ,118 | 35,218 | 1 | <0,001 | 2,009 | 1,596 | 2,530 |
| **Liver Disease** | ,851 | ,270 | 9,917 | 1 | ,002 | 2,342 | 1,379 | 3,977 |
| **Lung Disease** | ,471 | ,125 | 14,146 | 1 | <0,001 | 1,602 | 1,253 | 2,048 |
| **Neuromuscular Disease** | ,886 | ,095 | 87,857 | 1 | <0,001 | 2,425 | 2,015 | 2,919 |

**Table 7:** Odds Ratio for the association between the age, categories of comorbidity and outcome (hospitalization) in patients with COVID-19. The continuous value of Age was used instead of an age group. Being a continuous variable, an Odds Ratio of 1.045 implies that, as the Age variable increases by 1, the probability of the patient having the outcome increases by 4.5%. **B**, **S.E**, and **Wald** are the unstandardized regression weight, how much the unstandardized regression weight can vary by, and test statistic for the individual predictor variable, respectively.

|  | **B** | **S.E.** | **Wald** | **df** | **Sig.** | **Exp(B)** | **95% C.I** |  |
| --- | --- | --- | --- | --- | --- | --- | --- | --- |
|  |  |  |  |  |  |  | **Lower** | **Upper** |
| **Age** | ,044 | ,001 | 1721,450 | 1 | <0,001 | 1,045 | 1,043 | 1,047 |
| **Cancer** | ,916 | ,083 | 120,287 | 1 | <0,001 | 2,498 | 2,121 | 2,942 |
| **Cardiac Disorder** | ,526 | ,054 | 93,696 | 1 | <0,001 | 1,692 | 1,521 | 1,881 |
| **Diabetes** | ,719 | ,061 | 137,713 | 1 | <0,001 | 2,053 | 1,821 | 2,315 |
| **Hematologic Disease** | 1,002 | ,162 | 38,278 | 1 | <0,001 | 2,724 | 1,983 | 3,741 |
| **HIV** | 1,079 | ,176 | 37,594 | 1 | <0,001 | 2,941 | 2,083 | 4,152 |
| **Renal Disease** | 1,396 | ,099 | 198,500 | 1 | <0,001 | 4,041 | 3,327 | 4,907 |
| **Liver Disease** | 1,309 | ,184 | 50,593 | 1 | <0,001 | 3,704 | 2,582 | 5,314 |
| **Lung Disease** | ,794 | ,095 | 69,540 | 1 | <0,001 | 2,213 | 1,836 | 2,667 |
| **Neuromuscular Disease** | 1,323 | ,080 | 275,328 | 1 | <0,001 | 3,756 | 3,212 | 4,391 |

**Table 8:**
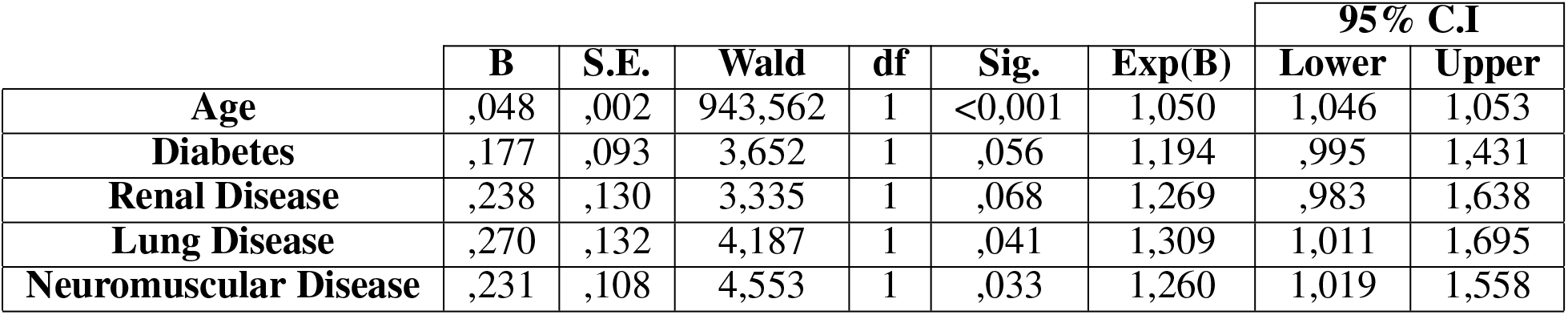
Odds Ratio for the association between the age, categories of comorbidity and outcome (ICU stay) in patients with COVID-19. The continuous value of Age was used instead of an age group. Being a continuous variable, an Odds Ratio of 1.050 implies that, as the Age variable increases by 1, the probability of the patient having the outcome increases by 5.0%. **B**, **S.E**, and **Wald** are the unstandardized regression weight, how much the unstandardized regression weight can vary by, and test statistic for the individual predictor variable, respectively.

**Table 9:** Odds Ratio for the association between the age, categories of comorbidity and outcome (Death + hospitalization + ICU stay) in patients with COVID-19. The continuous value of Age was used instead of an age group. Being a continuous variable, an Odds Ratio of 1.050 implies that, as the Age variable increases by 1, the probability of the patient having the outcome increases by 5.0%. **B**, **S.E**, and **Wald** are the unstandardized regression weight, how much the unstandardized regression weight can vary by, and test statistic for the individual predictor variable, respectively.

|  | **B** | **S.E.** | **Wald** | **df** | **Sig.** | **Exp(B)** | **95% C.I** |  |
| --- | --- | --- | --- | --- | --- | --- | --- | --- |
|  |  |  |  |  |  |  | **Lower** | **Upper** |
| **Age** | ,049 | ,001 | 2160,553 | 1 | <0,001 | 1,050 | 1,048 | 1,052 |
| **Cancer** | ,885 | ,084 | 111,355 | 1 | <0,001 | 2,423 | 2,055 | 2,855 |
| **Cardiac Disorder** | ,480 | ,054 | 78,761 | 1 | <0,001 | 1,616 | 1,454 | 1,797 |
| **Diabetes** | ,694 | ,061 | 127,705 | 1 | <0,001 | 2,002 | 1,775 | 2,258 |
| **Hematologic Disease** | ,983 | ,166 | 35,153 | 1 | <0,001 | 2,671 | 1,930 | 3,697 |
| **HIV** | 1,083 | ,176 | 37,806 | 1 | <0,001 | 2,955 | 2,092 | 4,174 |
| **Renal Disease** | 1,446 | ,102 | 201,261 | 1 | <0,001 | 4,246 | 3,477 | 5,184 |
| **Liver Disease** | 1,227 | ,186 | 43,534 | 1 | <0,001 | 3,409 | 2,368 | 4,908 |
| **Lung Disease** | ,830 | ,096 | 74,757 | 1 | <0,001 | 2,293 | 1,900 | 2,767 |
| **Neuromuscular Disease** | 1,375 | ,081 | 285,182 | 1 | <0,001 | 3,957 | 3,373 | 4,642 |

**Table 10:** Odds Ratio for the association between the age, number of comorbidities and outcome (Death + Hospitalization + ICU stay) in patients with COVID-19. Both Age and N^o^ of comorbidities are continuous variables, i.e, an Odds Ratio of 2.223 implies that, as the N^o^ of comorbidities increases by 1, the probability of the patient having the outcome increases by 122.3%. **B**, **S.E**, and **Wald** are the unstandardized regression weight, how much the unstandardized regression weight can vary by, and test statistic for the individual predictor variable, respectively.

|  | **B** | **S.E.** | **Wald** | **df** | **Sig.** | **Exp(B)** | **95% C.I** |  |
| --- | --- | --- | --- | --- | --- | --- | --- | --- |
|  |  |  |  |  |  |  | **Lower** | **Upper** |
| **Age** | ,050 | ,001 | 2321,879 | 1 | <0,001 | 1,051 | 1,049 | 1,053 |
| **N°of comorbidities** | ,799 | ,023 | 1257,322 | 1 | <0,001 | 2,223 | 2,127 | 2,323 |

When comparing the two versions of the dataset that were released by the DGS, it is possible to state that the June version is more complete and representative than the April one. This is easily verified by the prevalence of cardiac disorders in the total population, with a value of 0.28% in the April version and that rose to 8.14% in the most recent one (i.e., in the April version of the dataset all chronic diseases, including cardiac disorders, were encoded as a single categorical variable, and we believe that the information on cardiac disorders was also being extracted from textual notes by the DGS, given that the SINAVE system does not feature a structured field for collecting this information). Concerning chronic diseases and multimorbidity prevalence, the first version displayed values of 16.39% and 4.49%, respectively. With respect to Odd Ratios, the June version mainly showed differences in higher ratios for cardiac disorders. In the April version, due to the small sample size of patients with cardiac disorders, some Odd Ratios got as high as 91.72 (CI 95%: 28.61-294.06), as was the case for the composite outcome of death, hospitalization, or ICU admission (see Table 5).

As mentioned before, the way by which the structured information on chronic diseases was encoded changed amidst the two versions of the dataset. Besides the difference in terms of encoding the data as a single versus multiple categorical variables, the most recent version makes a distinction between *present, not present*, and *unknown*, while the April version listed the conditions that were *present* or *not present*. This difference can influence the veracity of the results, and we believe the more recent version can better depict the population affected by the different diseases and outcomes.

## 4 Discussion

Our study shows that multimorbidity is significantly associated with adverse outcomes for COVID-19 infection in the Portuguese population, independently from age. All chronic conditions, except asthma, lead to increased risk of hospitalization. However, only diabetes, chronic kidney disease, chronic respiratory diseases, and neuromuscular disorders, are associated with more severe cases requiring ICU admission. These results are in line with previous reports where chronic diseases are associated with poorer outcomes [3, 7]. Although the strength of association differs between diseases, every additional morbidity leads to an increased risk of the composite outcome of hospitalization, ICU admission, and mortality.

Multimorbidity was previously studied by Laires et al. for the general Portuguese population in 2014, using data from the fifth National Health Interview Survey (Inquérito National de Saude, INS) [6]. Prevalence of multimorbidity on the study from Laires at al. can be compared with our data. Although only individuals aged 25-79 were included in the study from 2014, the choice of individuals constitutes a robust sample for the study of morbidity prevalence in Portugal. We can observe in Figures 1 to 3 a rise in chronic health conditions with increasing age, as expected. However, multimorbidity is much less prevalent in our study population (6.77% vs 43.9%). Since the beginning of the COVID-19 pandemic there has been significant public awareness regarding the higher risk of older people with comorbidities. This may have induced efforts to protect and isolate this population, with the findings of our study suggesting a positive effect of such measures, given the younger and healthier population in the COVID-19 dataset. The discrepancy may however be related to different reporting methods. For instance, the maximum number of significant reported simultaneous morbidities in the COVID-19 infected population was 5 disorders (there were a total of 5 people with simultaneous morbidities ranging from 6 to 8), which is half of the maximum number of morbidities found in the INS population (10 disorders). Since the total number of conditions considered in both datasets is not so different (COVID-19: 10 diseases; INS: 13 diseases), a possible explanation for the higher number of co-occurring conditions in the INS population can be the combination of self-diagnoses with the presence of more *subjective* disorders, such as lower and upper back pain, allergies, depression, and urinary incontinence.

Our study also has several important limitations. First of all, the cross-sectional nature of the COVID-19 dataset makes it impossible to account for incomplete outcomes, since several patients could ultimately be hospitalized or die after the end of observation. Reported data on outcomes may therefore be underestimated, so careful interpretation is advised until more data is available. More importantly, despite the fact that no standard set of conditions is established to define multimorbidity, chronic conditions were given on broad groups and there is no specific information on individual conditions. For example, diabetes is given as one group and no distinction is made between type 1 and type 2 diabetes. Therefore, measured morbidities may herald heterogeneous groups of diseases with different degrees of severity, which may influence outcomes. Future datasets should ideally include more accurate information on chronic conditions.

Another important concern is related to the risk of under-reporting, which becomes obvious by analyzing reported cardiac diseases. Given that cardiovascular diseases, particularly hypertension, are very prevalent in the Portuguese population [8], the observed prevalence of 8.14% in our study highly suggests that under-reporting may have occurred. In addition, the prevalence of reported cardiac diseases has significantly increased from the April version of the DGS dataset, which showed a much lower cardiac disease prevalence in the general population of 0.28%. Surprisingly, cardiovascular diseases are absent from the available list of previous conditions in SlNAVE’s reporting page, which could have contributed to a lower notification of this comorbidity (see Figure 6b). One explanation to the considerable increase in cardiac disorder prevalence is the fact that the *raw* textual input from doctors was included in the June version of the DGS/SINAVE dataset and used in our analysis through the consideration of keywords, instead of relying on the categorical variable for the presence or absence of this disease, as in the case of the April version.

**Figure 5:**
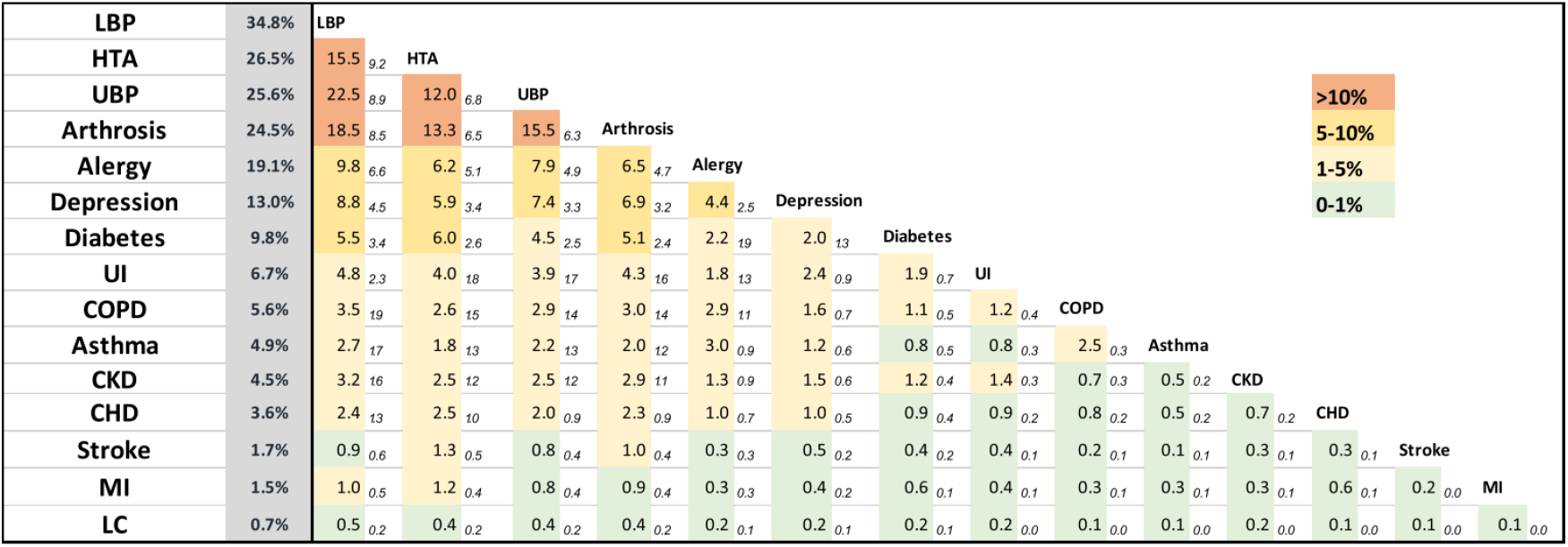
Percentage of observed and expected prevalence of co-occurring pairs of chronic health conditions (data from the Portuguese Fifth National Health Interview Survey) [6]. The shaded bar depicts the prevalence of each chronic health condition. In the matrix, the first value for each pair is the observed frequency, while the second (italic) is the expected one after multiplying the respective prevalence of each disorder. Chi-square tests were used to determine whether observed frequencies were significantly different from expected frequencies. All p-values were inferior to 5%.) LBP low back pain, HTA hypertension, UBP upper back pain, UI urinary incontinence, COPD chronic obstructive pulmonary disease, CKD chronic kidney disease, CHD coronary heart disease, MI previous myocardial infarction, and LC liver cirrhosis

**Figure 6:**
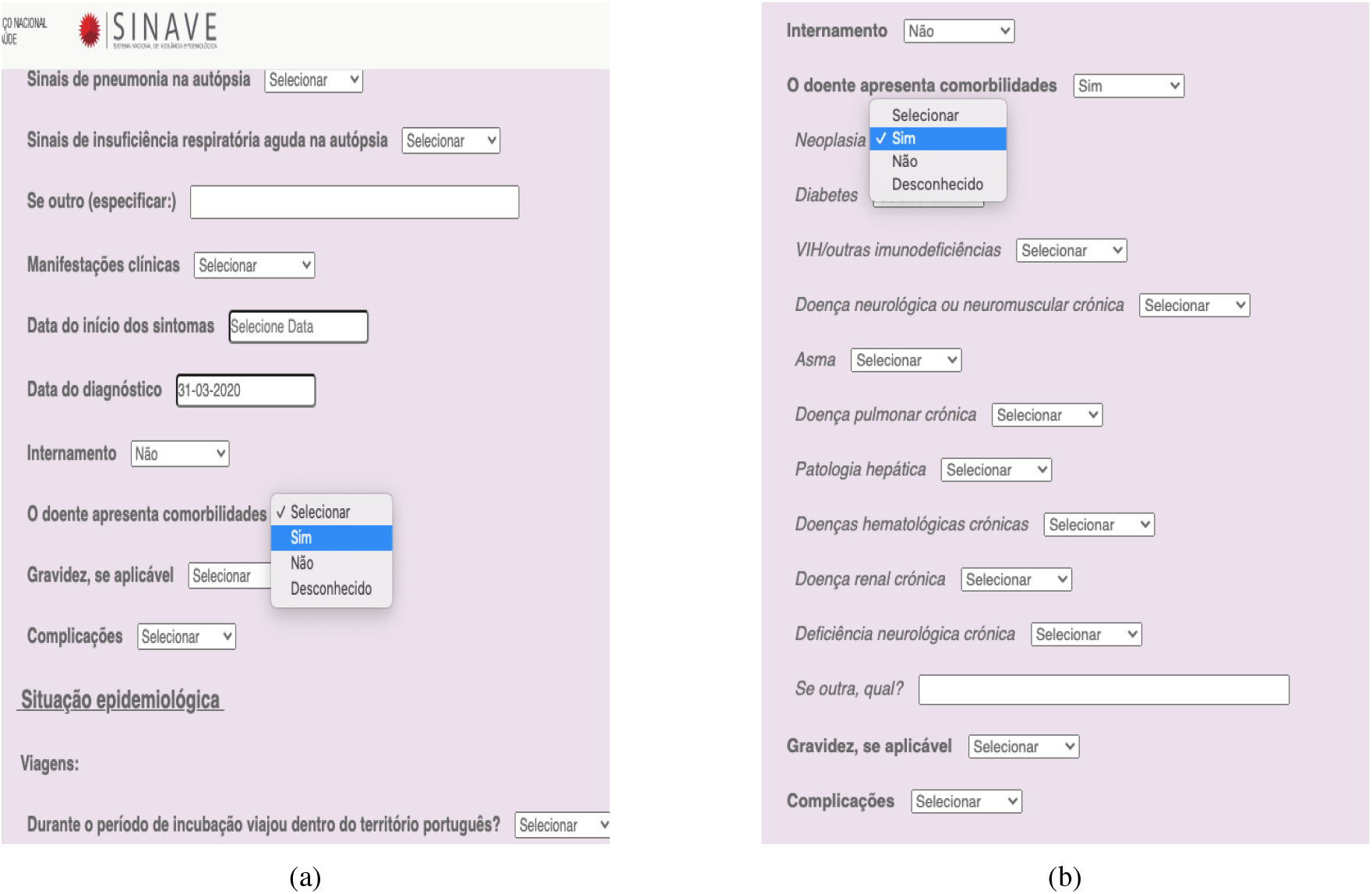
Screenshots of SINAVE’s interface regarding the report of known comorbidities: (a) Reporting the presence of comorbidities; (b) Reporting the known comorbidities

Although we acknowledge that the DGS/SINAVE dataset was not primarily generated for research, but rather for public health proceedings and government information, we believe that a better user interface design and a more rational set of chronic conditions could effortlessly improve the quality of the recorded data. One important lesson that we can learn from this pandemic is the important contribution that quickly gathered relevant clinical data through effective health information systems, such as SINAVE, can have in this setting.

We indeed suspect SINAVE to be very susceptible to under-reporting, specially regarding multimorbidity. The user interface is somewhat confusing and non-practical, allowing doctors to skip filling some important fields in the entry form. Some suggestions to improve future studies regarding multimorbidity, using SINAVE’s data, include:

- Redesign the reporting form, making data entry more effective and faster. The current user interface could be simplified, while encouraging the input of relevant comorbidity information
- Integrate SINAVE data with the patient’s health record or with data from the new *Trace COVID-19* system, to provide richer data relevant to COVID-19. This could be a way of implementing the redesigned user interface suggested above
- Make it mandatory to input if comorbidities are absent; if present, the filling of the entry form inputs related to comorbidities and the specific chronic conditions should be mandatory. As seen in Figure 6, the list of comorbidities only becomes visible if a previous parameter is filled
- Emphasize to healthcare professionals the importance of entering all known chronic conditions in the system
- Add cardiovascular diseases to the list of comorbidities.

To conclude, findings in our study show that multimorbidity is significantly associated with poor outcomes in COVID-19 infection. Further data is needed to inform about the strength of this association and about the significance of observed differences in multimorbidity prevalence between infected patients and the general population of Portugal. We believe that data collection problems may have occurred and influenced outcome measurement. We also provide recommendations for improving the data collection user interface, that could ultimately improve quality of health information about the COVID-19 infected population, while increasing confidence in SINAVE data.

## Data Availability

This retrospective observational study used data provided by DGS after the required institutional and ethical approvals. The sample population consists of all the Portuguese population with SARS-CoV2 confirmed infection as notified by clinicians by June 30, 2020. A broad range of clinical and demographic variables are present in this dataset. In this study we specifically used age, gender, hospital admission, admission in intensive care unit, mortality and patient's underlying conditions.

## Acknowledgements

This research was partially supported through the European Union’s Horizon 2020 research and innovation programme under grant agreement No 874850 (MOOD), as well as through the Fundação para a Ciência e Tecnologia (FCT) under the project grant with reference LISBOA-01-0247-FEDER-045948 (IntelligentCare), and through the INESC-ID multi-annual funding from the PIDDAC programme (UIDB/50021/2020).

1. List of Tables
2. Percentage of COVID-19 infected total Portuguese population affected by each comorbidity 11
3. Odds Ratio for the outcome (Death) for the Age variable and analyzed comorbidities. The real value of Age was used instead of an age group. Being a continuous variable, an Odds Ratio of 1.097 implies that, as the Age variable increases by 1, the probability of the patient having the outcome increases by 9 .7%. **B**, **S.E**, and **Wald** are the unstandardized regression weight, how much the unstandardized regression weight can vary by, and test statistic for the individual predictor variable, respectively. 12
4. Odds Ratio for the outcome (Hospitalization) for the Age variable and analyzed comorbidities. The real value of Age was used instead of an age group. Being a continuous variable, an Odds Ratio of 1.056 implies that, as the Age variable increases by 1, the probability of the patient having the outcome increases by 5.6%. **B**, **S.E**, and **Wald** are the unstandardized regression weight, how much the unstandardized regression weight can vary by, and test statistic for the individual predictor variable, respectively. 13
5. Odds Ratio for the outcome (ICU stay) for the Age variable and analyzed comorbidities. The real value of Age was used instead of an age group. Being a continuous variable, an Odds Ratio of 1.052 implies that, as the Age variable increases by 1, the probability of the patient having the outcome increases by 5.2. **B**, **S.E**, and **Wald** are the unstandardized regression weight, how much the unstandardized regression weight can vary by, and test statistic for the individual predictor variable, respectively. 14
6. Odds Ratio for the outcome (Death + hospitalization + ICU stay) for the Age variable and analyzed comorbidities. The real value of Age was used instead of an age group. Being a continuous variable, an Odds Ratio of 1.061 implies that, as the Age variable increases by 1, the probability of the patient having the outcome increases by 6.1%. **B**, **S.E**, and **Wald** are the unstandardized regression weight, how much the unstandardized regression weight can vary by, and test statistic for the individual predictor variable, respectively. 15
7. Odds Ratio for the association between the age, categories of comorbidity and outcome (Death) in patients with COVID-19. The continuous value of Age was used instead of an age group. Being a continuous variable, an Odds Ratio of 1.093 implies that, as the Age variable increases by 1, the probability of the patient having the outcome increases by 9.3%. **B**, **S.E**, and **Wald** are the unstandardized regression weight, how much the unstandardized regression weight can vary by, and test statistic for the individual predictor variable, respectively. 16
8. Odds Ratio for the association between the age, categories of comorbidity and outcome (hospitalization) in patients with COVID-19. The continuous value of Age was used instead of an age group. Being a continuous variable, an Odds Ratio of 1.045 implies that, as the Age variable increases by 1, the probability of the patient having the outcome increases by 4.5%. **B**, **S.E**, and **Wald** are the unstandardized regression weight, how much the unstandardized regression weight can vary by, and test statistic for the individual predictor variable, respectively. 17
9. Odds Ratio for the association between the age, categories of comorbidity and outcome (ICU stay) in patients with COVID-19. The continuous value of Age was used instead of an age group. Being a continuous variable, an Odds Ratio of 1.050 implies that, as the Age variable increases by 1, the probability of the patient having the outcome increases by 5.0%. **B**, **S.E**, and **Wald** are the unstandardized regression weight, how much the unstandardized regression weight can vary by, and test statistic for the individual predictor variable, respectively. 18
10. Odds Ratio for the association between the age, categories of comorbidity and outcome (Death + hospitalization + ICU stay) in patients with COVID-19. The continuous value of Age was used instead of an age group. Being a continuous variable, an Odds Ratio of 1.050 implies that, as the Age variable increases by 1, the probability of the patient having the outcome increases by 5.0%. **B**, **S.E**, and **Wald** are the unstandardized regression weight, how much the unstandardized regression weight can vary by, and test statistic for the individual predictor variable, respectively. 19
11. Odds Ratio for the association between the age, number of comorbidities and outcome (Death + Hospitalization + ICU stay) in patients with COVID-19. Both Age and N^o^ of comorbidities are continuous variables, i.e, an Odds Ratio of 2.223 implies that, as the N^o^ of comorbidities increases by 1, the probability of the patient having the outcome increases by 122.3%. **B**, **S.E**, and **Wald** are the unstandardized regression weight, how much the unstandardized regression weight can vary by, and test statistic for the individual predictor variable, respectively. 20

**Abbreviations**
SINAVE: Sistema Nacional de Vigilância Epidemiológica
COVID-19: Coronavirus Disease 2019
SARS-CoV-2: Severe Acute Respiratory Syndrome Coronavirus 2, Multimorbidity, Chronic Conditions

## Footnotes

2 https://covid19.min-saude.pt/disponibilizacao-de-dados/

